# Concentrations and immunostimulatory potential of circulating cell-free membrane-bound and membrane-unbound mitochondrial DNA in preeclampsia

**DOI:** 10.1101/2021.02.02.21250841

**Authors:** Spencer C. Cushen, Contessa A. Ricci, Jessica L. Bradshaw, Talisa Silzer, Alexandra Blessing, Jie Sun, Sabrina M. Scroggins, Mark K. Santillan, Donna A. Santillan, Nicole R. Phillips, Styliani Goulopoulou

## Abstract

Cell-free circulating mitochondrial DNA (CFCmtDNA) is a damage-associated molecular pattern (DAMP) that can activate Toll-like receptor 9 (TLR-9). The main objectives of this case-control study were 1) to determine absolute concentrations and immunostimulatory capacity of CFCmtDNA, in membrane-bound and -unbound states, in cases with preeclampsia and healthy controls and 2) to implement a bootstrapped penalized regression analysis to establish the contribution of CFCmtDNA to preeclampsia diagnosis and its interaction with commonly collected patient characteristics. To determine the contribution of membrane-bound and -unbound CFCmtDNA in preeclampsia, DNA from plasma samples was exctracted with lysis buffer (membrane-unbound) and without lysis buffer (membrane-bound). CFCmtDNA, quantified using absolute PCR quantification protocol, was reduced in preeclampsia compared to healthy controls (P≤0.02). While the pattern of reduced CFCmtDNA in preeclampsia was similar between methods of DNA extraction, DNA isolation with membrane lysis buffer resulted in 1,000-fold higher CFCmtDNA quantification in the preeclampsia group (P=0.0014) and 430-fold higher CFCmtDNA quantification in the control group (P<0.0001). Even though CFCmtDNA concentrations were reduced, plasma from women with preeclampsia induced greater TLR-9 activation than plasma from gestational age matched controls (P≤0.01) as monitored using SEAP reporter 293 cells expressing human *TLR-9*. Penalized regression analysis showed that women with preeclampsia are strongly likely to have high concentrations of nDNA and DNase I along with a prior history of preeclampsia. Low concentrations of CFCmtDNA and mode of delivery were also associated with preeclampsia. In conclusion, our data demonstrate increases in the immunostimulatory potential of CFCmtDNA and upregulation of DNA degradation mechanisms in women with preeclampsia at the third trimester.

## INTRODUCTION

Preeclampsia is a prevalent, multifactorial complication of pregnancy diagnosed as de novo hypertension after the 20^th^ week of gestation with end-organ damage^1^. This pregnancy-specific syndrome, which is seen in approximately 5-8% of pregnancies worldwide, results in increased morbidity and mortality for fetus and mother alike. Currently, there is no cure for preeclampsia and the most common approach to clinical management is delivery of the fetoplacental unit^2^, which often occurs preterm.

Often placentas from pregnancies with preeclampsia exhibit signs of damage in their functional structures, the placental villi^3^, and exaggerated rates of trophoblast cell death^4^. It has been proposed that placental damage results in release of anti-angiogenic factors and pro-inflammatory cytokines along with cellular constituents and DNA fragments, including mitochondrial DNA^5, 6^, into the maternal circulation. Thus, circulating factors are currently being studied as potential markers of early diagnosis and placental health, and as predictors of pregnancy outcomes^7-9^.

Cell-free circulating mtDNA (CFCmtDNA) can act as a damage-associated molecular pattern (DAMP) molecule, which elicits an immune response via activation of the pattern recognition receptor, Toll-like receptor-9 (TLR-9)^6^. We and others have previously reported that activation of TLR-9 during pregnancy induced preeclampsia-like signs in rats and mice^10, 11^. In line with these observations, TLR-9 activity was greater in pregnant women with preeclampsia as compared to healthy pregnant women^12^. Cross-sectional and case-control studies have reported increased CFCmtDNA copy number in serum and whole blood from women with preeclampsia in the third trimester^13, 14^ or at time of delivery^15^. On the other hand, CFCmtDNA was suppressed in the first trimester in women who later developed preeclampsia^16^. These previous studies suggest that CFCmtDNA may be a potential pathogenic trigger or a contributor to the maintenance of the maternal syndrome in preeclampsia.

The complex and multidimensional setting of preeclampsia poses many challenges when elucidating the role of CFCmtDNA in the pathophysiology of preeclampsia. Before determining whether CFCmtDNA is a trigger or a contributor to the maternal syndrome of preeclampsia, it is imperative that we first address these challenges, providing accurate and reliable quantifications. Thus, the main objectives of this case-control study are to: a) overcome the current challenges of measuring CFCmtDNA, particularly in limited sample sizes from pregnant women and b) determine the contribution of CFCmtDNA to preeclampsia diagnosis and its interaction with commonly collected patient characteristics.

One of the challenges of measuring CFCmtDNA is the accuracy of current protocols in determining concentrations of CFCmtDNA in biological samples. Most methods utilize relative PCR quantification approaches, which rely on a nuclear housekeeping gene. However, we have found that concentrations of nuclear DNA (nDNA) increase with advancing gestational age during pregnancy^17^, making quantification of CFCmtDNA relative to nDNA liable to produce spurious results. In the present study, we overcome this challenge by using our previously published absolute PCR quantification protocol that does not rely on measurements of a housekeeping gene^17^. A second challenge is that a significant part of mitochondrial genome is duplicated in the nuclear genome^18^ and therefore, it can be co-amplified by primers targeting CFCmtDNA. Here, we address this limitation selecting for amplification a mitochondria-specific genomic region that is absent from the nuclear genome^19, 20^. A third challenge is the appropriate implementation of multivariate statistical methods, which can be used to identify the relationship between circulating factors, such as CFCmtDNA, with other variables in the prediction of preeclampsia. Although logistic regression is often used when probing factors underlying disease occurrence, small sample sizes lead to overfitting even in balanced designs. To address this challenge, we employ bootstrapped penalized regression analysis, a robust approach that produces reliable and generalizable results^21, 22^. This method allows us to evaluate the contribution of DNA metrics to preeclampsia, in small sample sizes, while considering clinically significant covariates.

## METHODS

Detailed methods are available in the on-line Data Supplement.

### Subjects

De-identified subject information and blood samples were acquired from the Maternal Fetal Tissue Bank (IRB# 200910784) of the Women’s Health Tissue Repository at the University of Iowa^23^. Cases consisted of 19 pregnant women with preeclampsia and 19 healthy pregnant controls matched for gestational age (Table S1). As expected, the preeclampsia group exhibited significantly higher mean, systolic, and diastolic blood pressures and a significant lower gestational age of delivery, neonatal weight, and APGAR scores (P < 0.05) in comparison to the control pregnancies. Maternal venous blood was collected in the third trimester (mean = 33.6 weeks) and separated into plasma and peripheral blood mononuclear cells (PBMCs) before storing at -80°C until further processing.

### DNA quantification

DNA was isolated and quantified as published previously^17^, with a few modifications. Previous studies have noted that CFCmtDNA can be membrane-bound or -unbound^24, 25^, suggesting vesicular and non-vesicular transport of CFCmtDNA within the circulation. To determine the contribution of membrane-bound and -unbound CFCmtDNA in preeclampsia, DNA from plasma samples was isolated with lysis buffer and without lysis buffer. TaqMan™ chemistry-based absolute quantification of nDNA and CFCmtDNA was used as detailed elsewhere^17^. Isolated CFCmtDNA was quantified using a method modified from Kavlick et al.^20^ and detailed previously^17^. The target sequence for this analysis was the *MT-ND5* gene (mitochondrial NADH:ubiquinone oxidoreductase core subunit 5; GenBank Gene ID: 4540), spanning positions 13,288-12,392 of the mitochondrial genome^26^. The qPCR primers, probes, and synthetic DNA standards used for CFCmtDNA quantification are detailed in Table S2. Concentrations of plasma DNA are expressed as picograms (pg) per mL of plasma. Total DNA was calculated as the sum of CFCmtDNA and nDNA, in pg/mL plasma. Plasma CFCmtDNA is expressed in absolute concentrations as a percent of total DNA (%CFCmtDNA). The DNA content of PBMCs is presented as cell equivalent (Ceq) per microliter DNA isolate for nDNA and as CFCmtDNA genome copies per cell equivalent.

### Statistics

Normality tests, outlier removal and univariate analyses were performed using Prism (Version 8, GraphPad, San Diego, CA, USA). Student’s t-test and Mann-Whitney *U* test were used for group comparisons. The effect of fetal sex on primary outcomes was assessed with a two-way analysis of variance (ANOVA) followed by Sidak’s post-hoc analysis. DNA outcomes are presented in violin plots showing medians and ranges. Subject characteristics are presented as mean with minimum and maximum unless otherwise indicated. The significance level was set at *α* = 0.05 for all comparisons.

To identify the most important subject characteristics associated with the outcome of preeclampsia, bootstrapped penalized logistic regression was implemented. All elements of regression analyses were carried out in R software version 4.0.2^27-35^. Three penalized regression models (LASSO, ridge regression, elastic next regression) were fit and their performances compared to select the best model. To adhere to test assumptions of independent observations while not eliminating potentially important patient characteristics, related characteristics were grouped *a priori* (Table S3), and an in-house code was developed that selected a single characteristic from each group to generate all possible combinations of independent characteristics (*n* = 192 datasets). Bootstrapped (R = 500) 10-fold cross-validation LASSO, ridge, and elastic net were then performed on each dataset and best model fit was assigned by lowest prediction error. Predictive accuracy of the final model against a simulated naïve prediction dataset was assessed by the bootstrap estimate (R = 500) of the area under the curve of the receiver operator characteristic (AUC ROC) with confidence intervals (CI 95%) calculated from standard error. Characteristics selected by the final regression model at least 75% of the time (variable importance probability; VIP 0.75) for all bootstrap samples were regarded as most important^36^. Using this approach, the optimized predictor combination and model were thus selected to best explain the association between patient characteristics and the diagnosis of preeclampsia.

## RESULTS

Concentrations of CFCmtDNA in lysis buffer-treated samples (Figure 1A) and in samples that were not treated with lysis buffer (Figure 1B) were reduced in preeclampsia compared to healthy controls. While the pattern of reduced CFCmtDNA in preeclampsia remained the same between methods of DNA extraction, the exact quantity of CFCmtDNA was reduced by 1000-fold in samples never treated with lysis buffer (3.294 ± 0.828 vs. 0.003 ± 0.001 pg/mL, n=15, P=0.0014). In the control group, concentrations of CFCmtDNA were reduced by 430-fold in samples never treated with lysis buffer compared to samples treated with lysis buffer (9.011 ± 2.043 vs. 0.021 ± 0.004 pg/mL, n=18, P<0.0001). There were no group differences in plasma concentrations of nDNA in samples treated with lysis buffer (Figure 1C), while plasma concentrations of nDNA in samples treated without lysis buffer were increased in women with preeclampsia compared to healthy controls (Figure 1D). Content of CFCmtDNA and nDNA in PBMCs did not differ between groups (Figure 2A, B).

**Figure 1.**
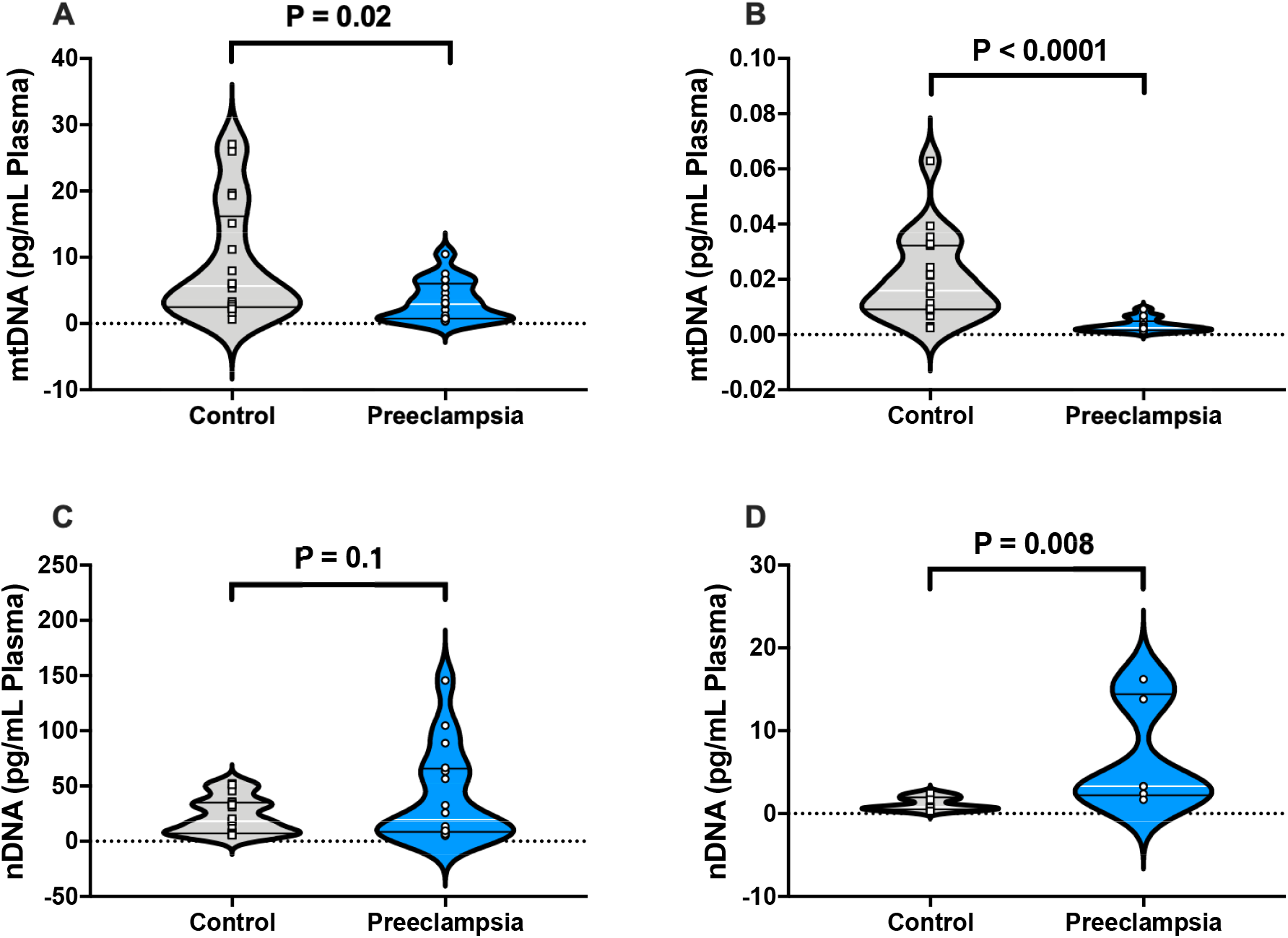
Absolute qPCR of DNA in maternal plasma. A) CFCmtDNA concentrations (pg/mL) in plasma isolated using lysis buffer, B) CFCmtDNA concentrations (pg/mL) in plasma isolated without lysis buffer, C) nDNA concentrations (pg/mL) in plasma isolated using lysis buffer, and D) nDNA concentrations (pg/mL) in plasma isolated without lysis buffer. A) and C) were analyzed using Student’s t-test while B) and D) were analyzed using Mann-Whitney *U* test. All values presented as median (white line) ± IQR (black lines). CFCmtDNA, cell-free circulating mitochondrial DNA; nDNA, nuclear DNA. Open squares = gestational age matched control; open circles = third trimester pregnancies with preeclampsia.

**Figure 2.**
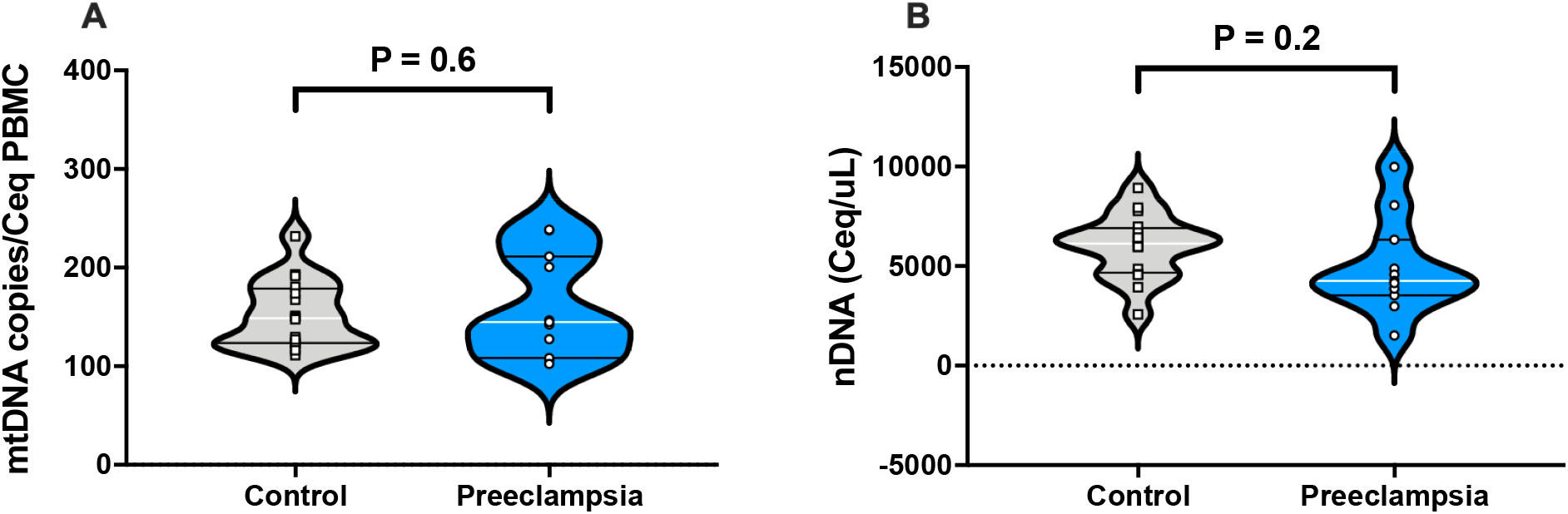
Absolute qPCR of DNA isolated from maternal peripheral blood mononuclear cells (PBMCs). A) mtDNA copies per estimated number of PBMC and B) nDNA concentration calculated as cell equivalents per microliter of PBMC DNA isolate. Data analyzed using Student’s t-test. All values presented as median (white line) ± IQR (black lines). nDNA, nuclear DNA; Ceq, cell equivalent. Open squares = gestational age matched control; open circles = third trimester pregnancies with preeclampsia.

Plasma DNA concentrations were compared between pregnancies carrying male and female fetuses. There was neither a main effect for sex nor a group-by-sex interaction for plasma CFCmtDNA concentrations (Figure 3A); however, there was an increase (P=0.0003) in circulating nDNA in pregnancies with preeclampsia carrying female fetuses (Figure 3B).

**Figure 3.**
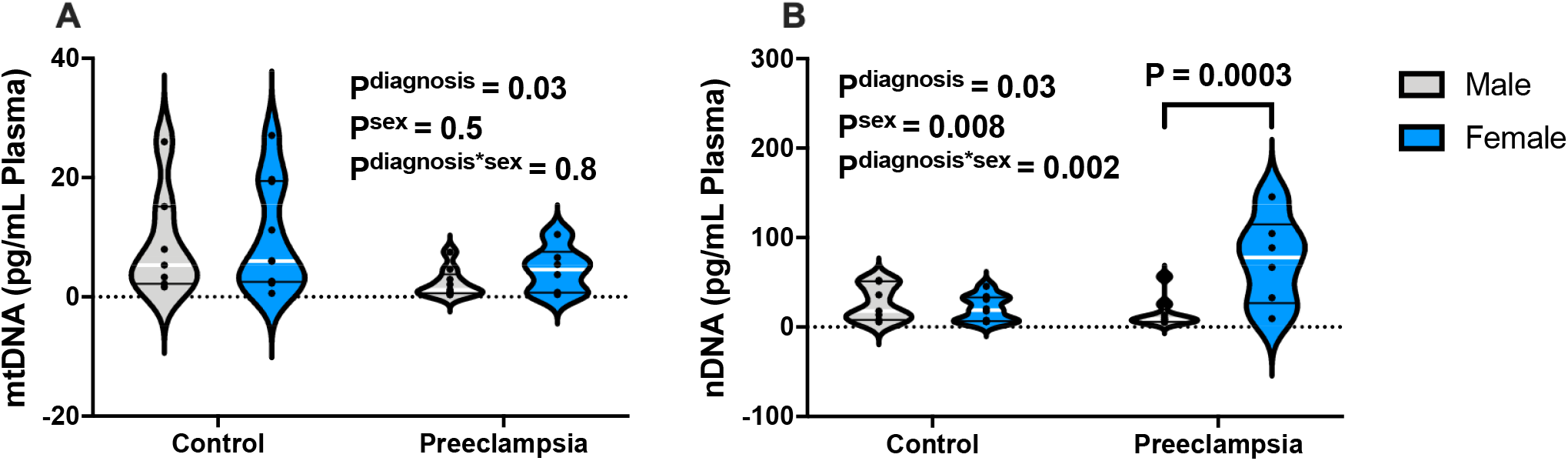
Effect of fetal sex on plasma concentrations of DNA. A) CFCmtDNA concentrations (pg/mL) in plasma and B) nDNA concentrations (pg/mL) in plasma. Data analyzed by two-way analysis of variance with Sidak’s post-hoc analysis. All values presented as median (white line) ± IQR (black lines). nDNA, nuclear DNA; CFCmtDNA, cell-free circulating mitochondrial DNA.

Even though CFCmtDNA absolute concentrations were reduced, plasma from women with preeclampsia induced greater TLR-9 activation than plasma from gestational age-matched controls, regardless of whether TLR-9 activity was normalized to lysis buffer extracted DNA (P=0.01, Figure 4A) or non-lysis buffer extracted DNA (P<0.0001, Figure 4B).

**Figure 4.**
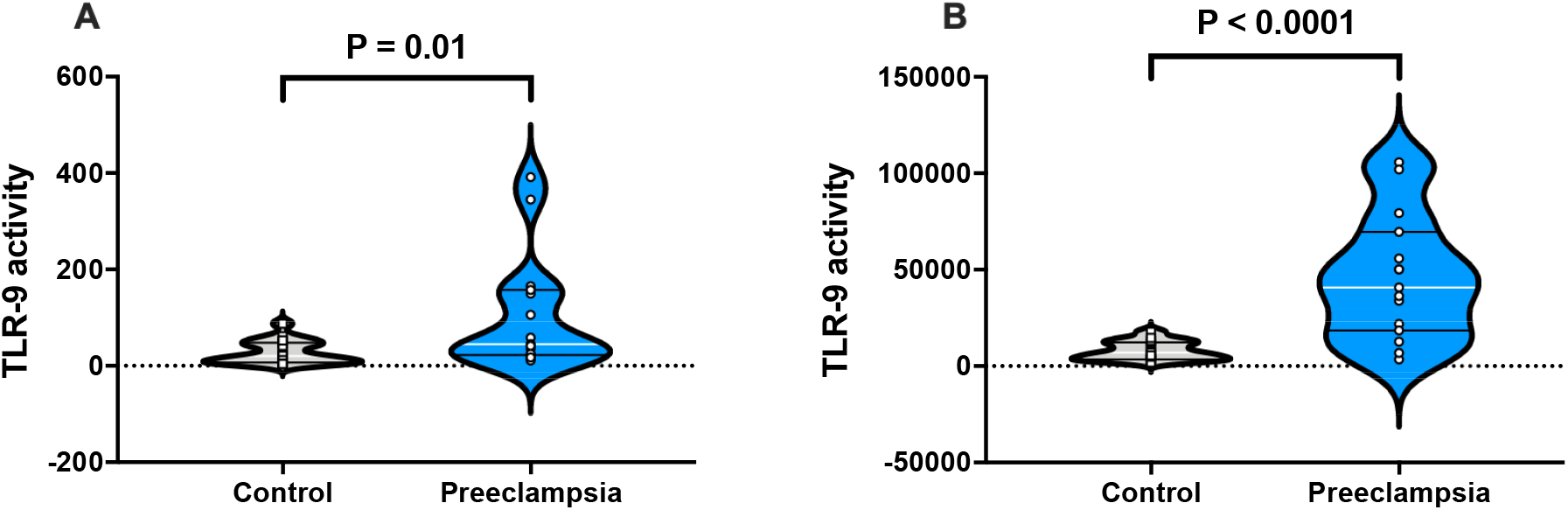
TLR-9 activity of SEAP reporter 293 cells exposed to maternal plasma. A) TLR-9 activity of cells exposed to 10% maternal plasma normalized to CFCmtDNA concentrations (pg/mL) in plasma when DNA was isolated using lysis buffer and B) TLR-9 activity of cells exposed to 10% maternal plasma normalized to CFCmtDNA concentrations (pg/mL) in plasma when DNA was isolated without lysis buffer. A) Mann-Whitney *U* test. B) Student’s t-test. All values presented as median (white line) ± IQR (black lines). TLR-9, toll-like receptor 9. Open squares = gestational age matched control; open circles = third trimester pregnancies with preeclampsia.

Univariate analysis showed no differences (P = 0.14) in plasma DNase I concentrations between control pregnancies and pregnancies with preeclampsia (Figure 5A). Moreover, there was no correlation (P=0.5) between DNase I concentrations and CFCmtDNA in either group (Figure 5B). Plasma DNase I was not correlated to nDNA (P=0.2) or %CFCmtDNA (P=0.3) in control pregnancies (Figure 5C, D). There were moderate correlations (P≤0.04) between DNase I and nDNA and %CFCmtDNA in preeclampsia (Figure 5C, D).

**Figure 5.**
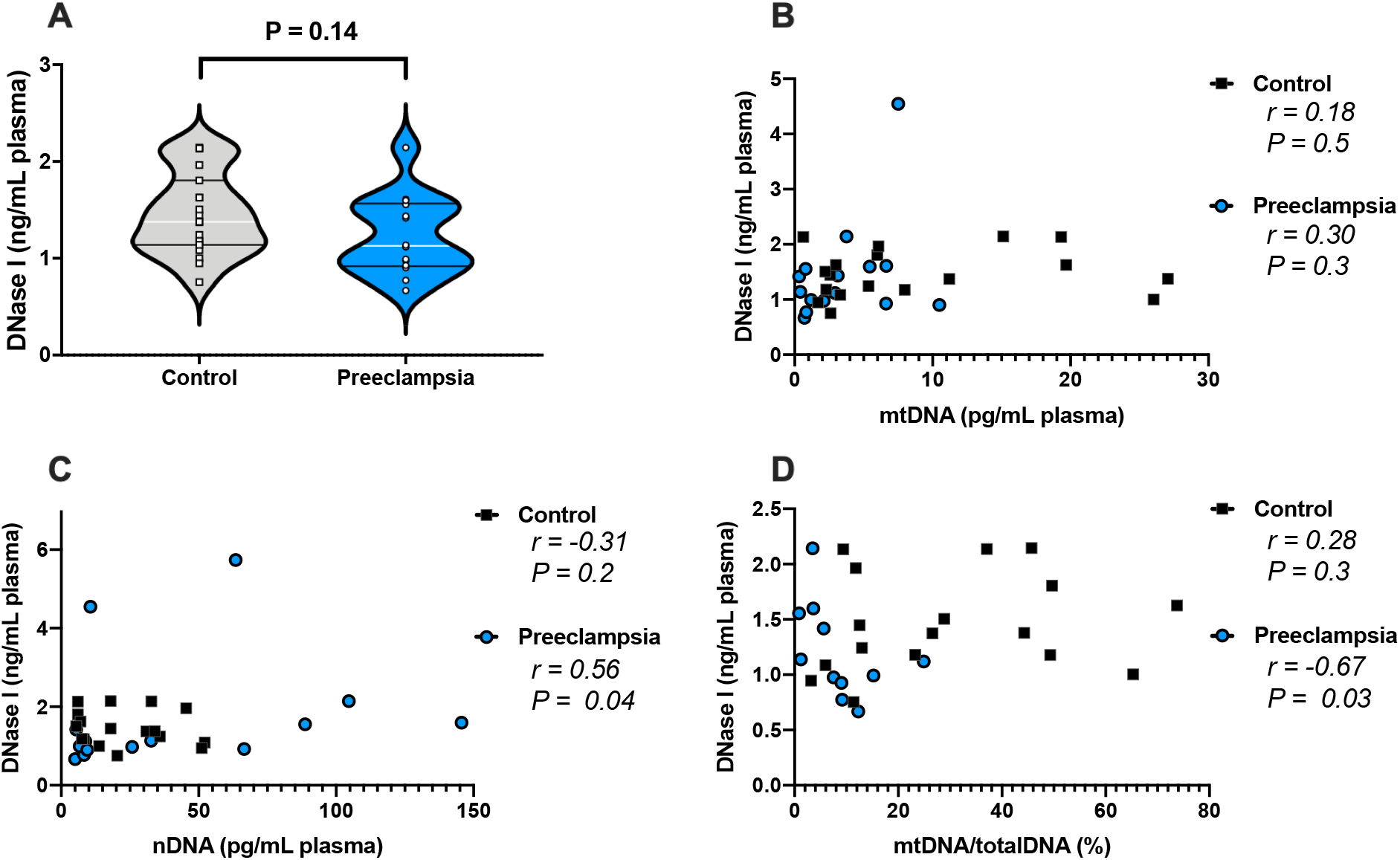
Plasma concentrations of DNase I measured in control pregnancies and pregnancies with preeclampsia. A) DNase I concentrations (ng/mL) in plasma, and DNase I plotted as a concentration against B) CFCmtDNA concentration (pg/mL) in plasma, C) nDNA concentration (pg/mL), and D) CFCmtDNA as a percent of total DNA. A) Student’s t-test, values presented as median (white line) ± IQR (black lines). B-D) Spearman correlation with r and P values presented for each analysis. CFCmtDNA and nDNA used for Spearman correlations were derived from samples treated with lysis buffer to extract DNA. nDNA, nuclear DNA; CFCmtDNA, mitochondrial DNA; totalDNA, sum of CFCmtDNA & nDNA. Open squares = gestational age matched control; open circles = third trimester pregnancies with preeclampsia.

A total of 192 datasets (all possible combinations of independent observations selected by in-house code in regression analysis) were analyzed to determine the best combination of variables for modeling the relationships between patient history and clinical characteristics, cell-free DNA, and the diagnosis of preeclampsia. The highest performing combination of characteristics and corresponding coefficients and odds ratios are listed in Table S4. After outlier removal via Mahalanobis distance, the final dataset used in model building included n=35 patients (preeclampsia, n=17; control, n=18). Assessing model performance between ridge, LASSO, and elastic net penalized regression of the best performing dataset over R=500 bootstraps showed that elastic net penalized regression produced the lowest prediction error (Table S5)^37, 38^. This model demonstrated an accuracy of approximately 100% (Table S6), and an AUC ROC value of approximately 1 (Table S6, Figure S1).

Variable importance probabilities at 75% (VIP 0.75) indicated that 9 of the subject characteristics considered were most important to the diagnosis of preeclampsia (Figure 6A). Characteristics with the largest effect (odds ratios) describing the diagnosis of preeclampsia were higher levels of nDNA and DNase I and having a history of preeclampsia (Figure 6B). Preeclampsia was also associated with lower levels of membrane unbound CFCmtDNA, higher likelihood of having a cesarean section, higher body mass index (BMI) at first obstetric visit, elevated mean arterial pressure and higher maternal age.

**Figure 6.**
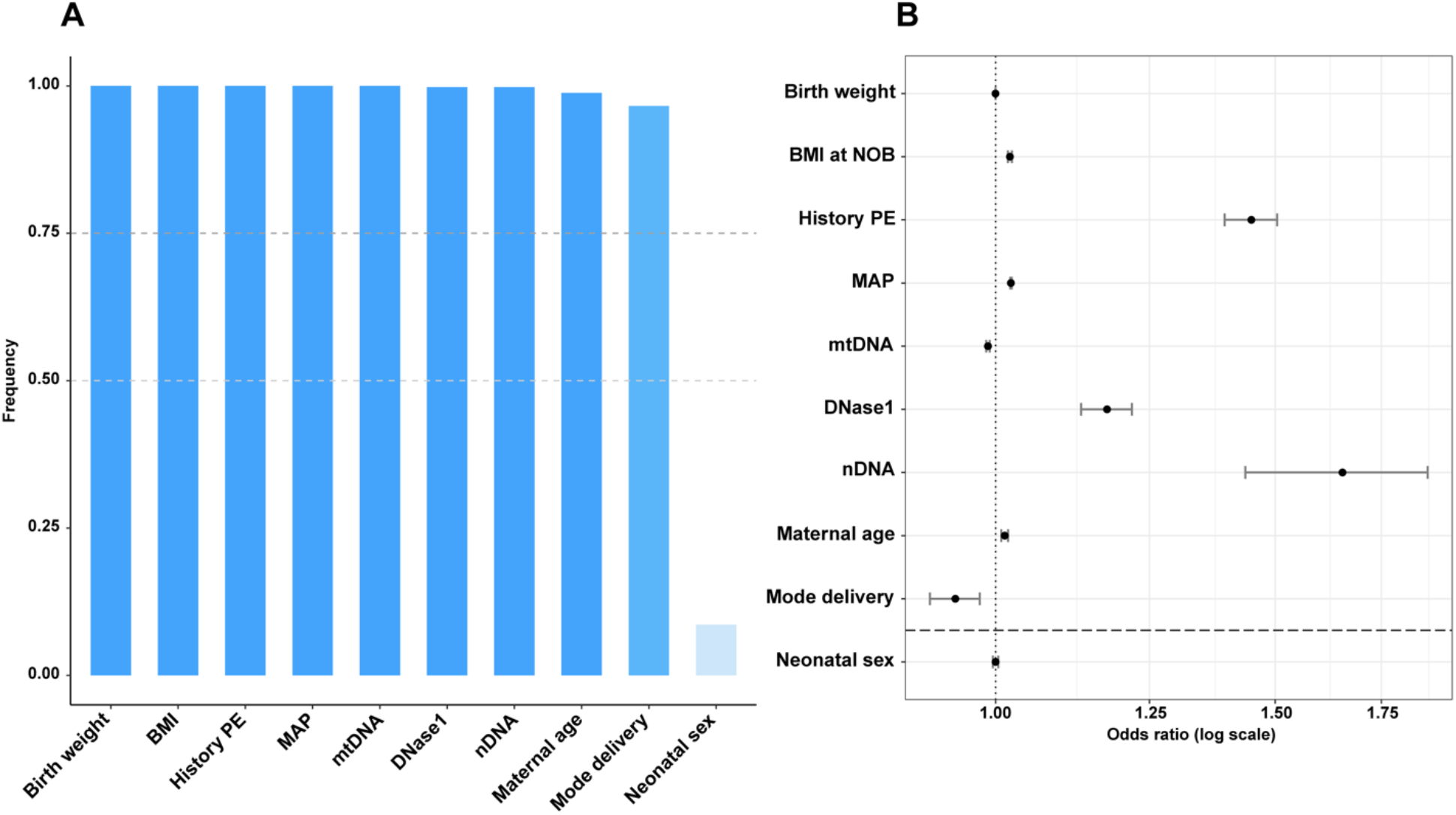
Subject characteristics most important to accurately determine preeclampsia via elastic net penalized regression. A) Subject characteristics whose variable importance probabilities (VIP) reached at least 75%. B) Odds ratios of selected subject characteristics. Vertical line represents OR = 1 (no effect); horizontal line represents VIP 0.75 cutoff.

## DISCUSSION

The main findings of this study are a) absolute concentrations of plasma CFCmtDNA are reduced in pregnant women with preeclampsia as compared to gestational age-matched controls at the third trimester; b) preeclampsia diagnosis is strongly associated with high circulating concentrations of nDNA, DNase I, and history of preeclampsia, and moderately associated with lower levels of membrane-unbound CFCmtDNA, higher maternal BMI, elevated mean arterial pressure, higher maternal age, and higher likelihood of a cesarean section. Additionally, our findings showed that the majority of CFCmtDNA within the maternal circulation is sequestered in vesicles during late pregnancy and this phenomenon is more pronounced in preeclampsia.

Our study demonstrates reduced CFCmtDNA in plasma from women with preeclampsia compared to controls. These quantities were determined in blood samples collected at the third trimester but before labor and delivery. In contrast, Marschalek *et al*. noted an increase in CFCmtDNA in serum of pregnant women (third trimester) suffering from preeclampsia when compared to controls^13^. Methodological differences in the protocol used to quantify CFCmtDNA may explain discrepancies between the results of this study and those from previous investigations. In our study, we aimed to address methodological limitations of previous research that might have introduced bias. We specifically chose to amplify a mitochondrial genome region (position 13,288-12,392) that has few known mutations and is absent from the nuclear genome. This approach protects against the effects of hypervariability and pseudogene contamination, respectively. To determine exact and not relative concentrations of CFCmtDNA, which would rely on a nuclear housekeeping gene, we used a double-stranded, synthetic, reference DNA. This method is highly sensitive for detecting minute quantities of CFCmtDNA^20^. Compared to these previous published protocols, these methodological changes improve accuracy and help ensure unbiased results.

Extracellular CFCmtDNA can elicit a pro-inflammatory response driven by activation of TLR-9^6, 39, 40^. TLR-9 specifically recognizes hypomethylated cytidine-phosphate-guanosine (CpG) motifs, which are common in bacterial DNA and mtDNA, but not in nuclear DNA^41^. In the context of our study, none of the study participants had an ongoing bacterial infection, and thus, we suggest that activation of TLR-9 was elicited by mtDNA. Although concentrations of CFCmtDNA was reduced in our cohort of women with preeclampsia, plasma from these subjects induced greater TLR-9 activity as monitored using SEAP reporter 293 cells expressing the human *TLR-9* gene. This finding is in agreement with previous studies showing increased TLR-9 activity in women with preeclampsia at time of diagnosis^12^. Our study extends these findings demonstrating greater TLR-9 activity in the presence of lower concentrations of CFCmtDNA in plasma from women with preeclampsia. This result suggests that CFCmtDNA in plasma from women with preeclampsia has greater immunostimulatory potency as compared to CFCmtDNA from normal pregnant women. Previous studies have shown that CFCmtDNA modifications, such as oxidation, increase the immunostimulatory potential of CFCmtDNA^6^. It is unknown whether CFCmtDNA of women with preeclampsia undergoes modifications that alter their immunostimulatory capacity.

While the pattern of reduced CFCmtDNA in preeclampsia was similar between methods of DNA extraction, DNA isolation with membrane lysis buffer resulted in 1,000-fold higher CFCmtDNA quantification in the preeclampsia group and 430-fold higher CFCmtDNA quantification in the control group. Membrane-bound and -unbound nDNA was also determined in these samples and although the membrane-unbound fraction was greater than the membrane-bound nDNA, the magnitude of increase was much smaller than what we observed for CFCmtDNA (Control: 27-fold increase, Preeclampsia: 15-fold increase). Thus, most of the cell-free DNA, and particularly CFCmtDNA, is membrane-bound in the maternal circulation in normal pregnancy and more so in preeclampsia. These results are in agreement with previous studies reporting that although circulating placenta-derived extracellular vesicles are present in both normal pregnancy and in preeclampsia, the concentration of these vesicles is greater in preeclampsia^42^. While mtDNA-containing extracellular vesicles have not been identified in the context of preeclampsia, exosomes and mitochondria-derived vesicles have been identified to contain CFCmtDNA^43^. Greater cell-free, yet membrane-bound, CFCmtDNA could be a component of several extracellular vesicles, including apoptotic bodies^43^, which may be related to changes in placental apoptosis in preeclampsia^44-48^.

In this study, we did not interrogate the source of CFCmtDNA. While it has been demonstrated that circulating cell-free DNA is associated with placental size and apoptosis of trophoblasts^49^, it has yet to be shown whether the same relationship exists between CFCmtDNA and placental dysfunction. Interestingly, placentas from preeclamptic pregnancies are smaller and experience reduced perfusion^50-54^ and have decreased villous surface area, suggesting that there is less area for maternal-fetal communication, even when adequate perfusion is present^55-58^. Placental apoptosis is increased^44-48^ and mitochondrial size, content, and bioenergetics are also altered in preeclampsia^44, 59-61^, and these changes may be secondary to placental hypoxia and oxidative stress ^44, 59, 60^. Whether placental dysfunction in preeclampsia contributes to CFCmtDNA and whether CFCmtDNA concentrations in the circulation reflect mitochondrial health in the placenta warrants further investigation.

Previous studies have suggested that plasma CFCmtDNA and cellular mtDNA copy number reflect different cellular processes in disease conditions. Intracellular mtDNA copy number in PBMCs is thought to reflect variations in mitochondrial bioenergetics and biogenesis^62^, while CFCmtDNA is mostly associated with cellular stress and tissue damage^6^. Here, in addition to plasma CFCmtDNA concentrations, we measured mtDNA copy number in PBMCs. In contrast to our findings in plasma samples, we detected no group differences in PBMC mtDNA copy number, suggesting dysregulation only occurred for extracellular mtDNA in this cohort of women with preeclampsia.

Greater circulating nDNA, but not CFCmtDNA, was found in pregnant women with preeclampsia who were carrying female fetuses than women carrying male fetuses. This relationship was not observed in the control group. Female fetal sex is associated with increased early preeclampsia and villous infarction^63^, and male sex is associated with term preeclampsia^64^. In the context of preeclampsia, the placentas from female offspring had greater villous infarction^65^, while placentas from male offspring had greater apoptosis and inflammation^66^. Greater cell-free circulating nDNA in pregnancies carrying female fetuses may reflect a sex-specific placental response in preeclampsia that can be assessed in the maternal blood. It is noteworthy that our study was not powered to determine the effects of fetal sex on cell-free DNA quantifications. Thus, we should interpret these findings with caution.

We then examined the relationship between maternal and neonatal characteristics and cell-free DNA by optimizing for the most informative subject data and analyzing the magnitude of association and/or contribution to the diagnosis of preeclampsia using elastic net penalized regression. Previously known relationships, such as a higher likelihood of experiencing preeclampsia if a subject has a history of preeclampsia^67, 68^, and higher mean arterial blood pressure as part of the diagnostic criteria^1^, were corroborated by the elastic net regression model. Additionally, elastic net regression mirrored univariate analyses showing lower levels of membrane-unbound CFCmtDNA associated with preeclampsia. We also found that higher concentrations of nDNA and DNase I are significant contributors to the diagnosis of preeclampsia. These results contrast the outcome of DNase I in univariate tests. Although plasma concentrations of DNase I were not significantly different in univariate tests, when considered with patient medical histories, clinical data, and cell-free DNA metrics, higher levels of DNase I may be representative of important processes that might otherwise be missed by other analytical approaches. DNase I is a significant component of cell-free DNA degradation and DNase I activity has been shown to be increased in pregnant women with intrauterine growth restriction^69^, a common feature of pregnancies with preeclampsia. Collectively, we have demonstrated the utility of multivariate analyses in capturing a more comprehensive understanding of available data, as univariate testing may not reflect the importance that a variable possesses in a full model. This computational approach allows us to identify phenotypic variables that may contribute to preeclampsia.

Limitations of this study include a lack of diversity in subject race and ethnicity and small sample sizes, which may limit the generalizability of our results. Nonetheless, the demographics of the included subjects are representative of the Iowa population, where the samples were collected. Further, DNA degradation mechanisms were not studied extensively. Factors other than DNase I likely contribute to elimination of circulating cell-free DNA, including filtration by the kidney or spleen, differential flux in DNA release and uptake by tissues, and degradation by other DNA hydrolases^70^. Further assessment of placental function coupled with CFCmtDNA quantification could shed insight on the origins of CFCmtDNA. Despite these limitations, our study provided a novel, rigorous, and accurate measurement of exact concentrations of CFCmtDNA and assessed the immunostimulatory potential of this CFCmtDNA in women with preeclampsia. Finally, penalized regression analysis allowed for further interpretation of clinical and molecular data, as it would have been easy to overlook the contribution of DNase I concentrations when we found no difference in univariate analysis.

In conclusion, this study demonstrated that despite reductions in absolute concentrations of CFCmtDNA in late pregnancy, plasma from women with preeclampsia elicits greater TLR-9 activation compared to plasma from healthy pregnant women. Through the use of penalized logistic regression, we have also identified greater DNase I as a risk factor for preeclampsia.

## Data Availability

The data that support the findings of this study are available on request from the corresponding author.

## PERSPECTIVES

This study provides a new approach for assessing the role of CFCmtDNA in preeclampsia, implementing absolute quantification methods, amplifying a mitochondrial genomic region that is absent from the nuclear genome, and utilizing a penalized regression analysis to determine the contribution of circulating DNA and DNA degradation mechanisms to preeclampsia. This computational analysis is particularly robust for small sample sizes. Our data demonstrate increases in the immunostimulatory potential of CFCmtDNA and upregulation of DNA degradation mechanisms in women with preeclampsia during the third trimester. Additionally, this study sets the foundation for future investigations in the exact role of CFCmtDNA in the pathogenesis of preeclampsia, as previous studies have suggested that CFCmtDNA may be a potential pathogenic trigger or a contributor to the maintenance of the maternal syndrome in preeclampsia^13, 15, 16^.

## ACKNOWLEDGMENTS

Computational and Analytics resources were provided by the North Texas Scientific Computing, a division under the of office of the Chief Information Officer for UNT and UNT System. We would like to acknowledge Dr. Ravi Vadapalli and Dr. Richard Herrington (North Texas Scientific Computing) for reviewing R scripts and consulting on R code implementation.

## NOVELTY AND SIGNIFICANCE

### 1) What Is New

- Cell-free circulating mtDNA (CFCmtDNA) concentrations are reduced in plasma from women with preeclampsia at the third trimester.
- Despite having CFCmtDNA, plasma from women with preeclampsia elicits greater activation of TLR-9.
- Preeclampsia diagnosis is strongly associated with high circulating concentrations of nDNA, DNase I, and history of preeclampsia, and moderately associated with lower levels of membrane unbound CFCmtDNA, higher maternal BMI, elevated mean arterial pressure, higher maternal age, and higher likelihood of a cesarean section.

### 2) What Is Relevant?

- Preeclampsia is associated with placental dysfunction, trophoblast cell death, and oxidative stress, which lead to release of pro-inflammatory factors into the maternal circulation.
- An increase in the immunostimulatory potency of CFCmtDNA may contribute to the development or establishment of the maternal syndrome in preeclampsia.

