## Supplemental Materials for "Concentrations and immunostimulatory potential of circulating cell-free membrane-bound and membrane-unbound mitochondrial DNA in preeclampsia"

### Detailed Methods

#### Subjects and Experimental Design

This is a cross-sectional, case-control study. De-identified samples and subject information were acquired from the Maternal Fetal Tissue Bank of the Women's Health Tissue Repository at the University of Iowa[1]. Tissue bank inclusion and exclusion criteria have been previously published[1]. The Maternal Fetal Tissue Bank has been approved by the Institutional Review Board of the University of Iowa (IRB#200910784). Details about sample collection for the tissue bank are published elsewhere[1]. The present study was reviewed by the Institutional Review Board of the University of North Texas Health Science Center, which determined the protocol to meet criteria for exempt status (IRB#2017-065, exempt category 4).

Cases consisted of 19 pregnant women clinically diagnosed with preeclampsia. Nineteen healthy pregnant controls were matched to cases for gestational age at sampling. Subject characteristics can be found in Table S1. Maternal blood was collected in the third trimester (28-41 weeks of gestation) during routine venipuncture. Blood was collected into ACD-A tubes (Becton Dickinson) containing: 22.0 g/L trisodium citrate, 8.0 g/L citric acid, and 24.5 g/L dextrose, and stored in 4°C until further processing. Blood was then separated into plasma and peripheral blood mononuclear cells (PBMCs). PBMCs were stored in cryopreservation media (RPMI media [40% v/v], FBS [50%], and DMSO [10%]). Plasma samples were snap frozen and stored at -80°C and PBMCs were snap frozen and maintained in liquid nitrogen.

**DNA Measurements – Absolute quantification polymerase chain reaction (qPCR)**

DNA was isolated and quantified as published previously[2], with a few modifications. Briefly, DNA from plasma and PBMCs (200 µL) was isolated using a magnetic bead-based extraction method (Omega Bio-tek) with a final elution volume of 360 µL. DNA from plasma samples was isolated in the presence and absence of lysis buffer to elucidate the contribution of the membrane bound component of plasma, as this has been noted previously to contain mtDNA[3, 4].

TaqMan™ chemistry-based absolute quantification of nuclear DNA (nDNA) and mtDNA are detailed elsewhere[2]. Briefly, isolated nDNA (2 µL) was quantified on a 7500 Real-Time PCR System (Applied Biosystems) using the Quantifiler™ Trio DNA Quantification Kit (Applied Biosystems, Waltham, MA, USA; Cat. No. 4482910) in 18 µL of master-mix for a total reaction volume of 20 µL. PCR settings were as follows: 95 °C for 2 minutes and 40 cycles of 95 °C for 9 seconds with 60°C at 30 seconds. Cycle threshold ($C_T$) was compared to five 1:10 serial dilutions of male, genomic reference DNA in order to calculate a concentration [$nDNA/\mu L_{DNA\ isolate}$] according to the manufacturer's directions.

Isolated mtDNA was quantified using a method modified from Kavlick et al.[5] and detailed previously[2]. The target sequence for this analysis is the *MT-ND5* gene (mitochondrial NADH:ubiquinone oxidoreductase core subunit 5; GenBank Gene ID: 4540), spanning positions 13,288-12,392 of the mitochondrial genome (based on revised Cambridge Reference Sequence positions)[6]. Isolated mtDNA (2 µL) was added to 23 µL of master-mix for a total reaction volume of 25 µL. Quantification was performed on a 7500 Real-Time PCR System (Applied Biosystems), with the following settings: 9600 emulation, 50 °C for 2 minutes, 95 °C for 10 minutes, and 40 cycles of 95 °C for 15 seconds with 60 °C for 1 minute. $C_T$ of samples was compared to eight, 1:10, serial dilutions of double-stranded, synthetic, reference DNA (gBlocks® gene fragment; Integrated DNA Technologies, Coralville, IA, USA) in order to calculate a concentration of mtDNA in the isolate [mtDNA/$\mu L_{DNA\ isolate}$]. The qPCR primers, probes, and synthetic DNA standards employed for mtDNA analysis are detailed in Table S2.

For DNA quantification, amplification efficiency >80% and $R^2$ >99% was considered adequate. Concentration of plasma DNA ($[DNA]^{plasma}$) was determined by relating the calculated concentration of the DNA lysate ($[DNA]^{isolate}$) multiplied by the volume of elution buffer (0.360 mL) to the known volume of isolated plasma (0.200 mL) (equation 1a-b), and it was expressed as picograms (pg) per mL of plasma. Total DNA was calculated as the sum of mtDNA and nDNA, in pg/mL plasma.

$$[DNA]^{plasma}\ pg/mL \times\ 0.2\ \text{mL}\ =\ [DNA]^{isolate} pg/mL\ \times\ 0.36\ \text{mL} \qquad \text{(1a)}$$

$$[DNA]^{plasma}\ pg/mL\ =\ \frac{0.36\ \text{mL} \times [DNA]^{isolate}\ pg/mL}{0.2\ \text{mL}} \qquad \text{(1b)}$$

The DNA content of PBMCs is presented as cellular equivalents (Ceq) per microliter DNA isolate [pg/$\mu L_{DNA\ isolate}$] for nDNA and as mtDNA genome copies per Ceq. Ceq was calculated based on the estimated molecular weight of nDNA per human diploid cell of 6.7pg (equation 2a). mtDNA copies were calculated by relating number of mitochondrial genomes that has a mass of 1 pg, where 1 pg is equal to 58,800 mitochondrial genome copies (equation 2b).

$$Ceq^{PBMC}/\mu L\ =\ [nDNA]^{PBMC}\ pg/\mu L\ \times\ \frac{1\ Ceq}{6.7\ pg} \qquad \text{(2a)}$$

$$[mtDNA]^{PBMC}\ copies/\mu L\ =\ [mtDNA]^{PBMC}\ pg/\mu L\ \times\ \frac{58{,}800\ copies}{1\ pg} \qquad \text{(2b)}$$

**DNase I measurement in maternal plasma**

DNase I concentrations in maternal plasma were measured using an ELISA (MyBioSource, Ca MBS763541). Plasma samples were diluted 1:10 before performing the ELISA per manufacturer's instructions.

**TLR-9 stimulation**

To determine the immunostimulatory potency of plasma from pregnancies with preeclampsia in relation to TLR-9 activation, we used an engineered cell line of human embryonic kidney (HEK) 293 cells transfected with a human *TLR-9* gene (HEK-Blue$^{TM}$ hTLR-9 cells, Invivogen). HEK-Blue$^{TM}$ hTLR-9 cells express an inducible secreted embryonic alkaline phosphatase (SEAP) reporter gene under the control of a fused promoter containing NF-κB and AP-1 binding sites. Stimulation of HEK-Blue$^{TM}$ hTLR-9 cells with a TLR-9 ligand activates NF-κB and AP-1, which induce the production of SEAP. HEK-Blue$^{TM}$ hTLR-9 cells were cultured and maintained according to the manufacturer's instructions. TLR-9 stimulation was assessed in response to 10% plasma from women with healthy pregnancies or pregnancies complicated by preeclampsia by monitoring SEAP production in a cell-culture detection medium (HEK-Blue$^{TM}$ Detection, Invivogen). TLR-9 agonists (ODN 2006 and ODN 2395, Invivogen) and antagonist (ODN 2088, Invivogen) were used as positive and negative controls of SEAP production, respectively. After 24 hours of incubation, SEAP production was quantified by reading the optical density of samples at 630 nm using a BioTEK Synergy HTX spectrophotometer.

**Data Analysis and Statistics**

To identify the most important patient characteristics associated with the outcome of preeclampsia, bootstrapped penalized logistic regression was implemented. All elements of regression analyses were carried out in R software version 4.0.2[7]. The following penalized regression models were fit and their performances compared to select the best model: 1) least absolute shrinkage and selection operator (LASSO) regression; 2) ridge regression; and 3) elastic net regression. These models perform optimally under different conditions: LASSO performs best when few predictors influence the outcome[8]; ridge performs best when many predictors have a small effect on the outcome[8, 9]; and elastic net performs best when the dataset is an intermediate between the two[10]. To adhere to test assumptions of independent observations while not eliminating potentially important patient characteristics, related characteristics were grouped *a priori* (Table S3) and an in-house code was developed that selected a single characteristic from each group to generate all possible combinations of independent characteristics ($n$ = 192 datasets). Bootstrapped (R = 500) 10-fold cross-validation LASSO, ridge, and elastic net were then performed on each dataset and best model fit was assigned by lowest prediction error (specifically, lowest median root mean squared error; RMSE[11, 12]). Predictive accuracy of the final model against a simulated naïve prediction dataset was assessed by the bootstrap estimate (R = 500) of the area under the curve of the receiver operator characteristic (AUC ROC) with confidence intervals (CI 95%) calculated from standard error. Characteristics selected by the model at least 75% of the time (variable importance probability; VIP 0.75)[13] for all bootstrap samples were regarded as most important. Using this approach, the optimized predictor combination and model were thus selected to best explain the association between patient characteristics and the diagnosis of preeclampsia.

<u>Statistics</u>

Statistical analyses were performed using Prism (Version 8, GraphPad, San Diego, CA, USA). Data distribution was assessed using the D' Agostino-Pearson omnibus test and the robust regression and outlier removal (ROUT) method was used to identify and remove outliers. Non-parametric statistics were used for non-normally distributed data sets. Group differences in DNA quantities and DNase I concentrations were determined using Student's t-test or Mann-Whitney *U* test. A two-way analysis of variance (ANOVA) followed by Sidak's post-hoc analysis was used determine the effect of fetal sex on group differences in DNA quantities. Spearman correlation was applied to evaluate relationships between DNA quantities and DNase I for each group. DNA outcomes are presented as mean $\pm$ standard error of the mean (SEM). Subject characteristics are presented as mean with minimum and maximum unless otherwise indicated. Exact P values are presented for each analysis.

**<u>Supplementary Tables</u>**

**Table S1. Subject characteristics**

| | *Control* (n = 19) | *Preeclampsia* (n = 19) | *P value* |
|---|---|---|---|
| *Race (%)* | | | |
| Caucasian | 19 (100) | 18 (95) | |
| Non-Hispanic | 19 (100) | 19 (100) | |
| *BMI (kg/m$^2$)* | 27 (18, 46) | 31 (18, 50) | 0.1 |
| Overweight (BMI: 25-29.9) (%) | 3 (16) | 7 (37) | |
| Obese (BMI: ≥ 30) (%) | 4 (21) | 8 (42) | |
| *MAP (mmHg)* | 90 (76, 101) | 108 (86, 129) | <0.0001 |
| SBP (mmHg) | 124 (105, 140) | 146 (122, 170) | <0.0001 |
| DBP (mmHg) | 73 (57, 87) | 89 (67, 112) | <0.0001 |
| *History of chronic hypertension (%)* | 0 (0) | 8 (42) | |
| *History of preeclampsia (%)* | 0 (0) | 4 (21) | |
| *Medications (%)* | | | |
| Aspirin | 0 (0) | 3 (16) | |
| Magnesium | 0 (0) | 3 (16) | |
| Nifedipine | 0 (0) | 2 (11) | |
| *Gestational age at sample (weeks)* | 33 (28, 39) | 34 (28, 41) | 0.7 |
| *Gestational age delivery (weeks)* | 39 (37, 42) | 36 (31, 41) | 0.0002 |
| *Mode of delivery (%)* | | | |
| NSVD | 14 (74) | 8 (42) | |

| | | | |
|---|---|---|---|
| VAVD | 0 (0) | 2 (11) | |
| Cesarean section | 5 (26) | 9 (47) | |
| *Neonatal weight (kg)* | 3.3 (1.5, 4.6) | 2.7 (1.5, 3.9) | 0.0076 |
| *Neonatal sex (F:M)* | (12:7) | (7:12) | |
| *Apgar, 1 minute* | 7.58 (3,9) | 7.05 (4, 9) | 0.4 |
| *Apgar, 5 minutes* | 8.95 (8, 9) | 8.42 (6, 9) | 0.028 |

Maternal BMI, MAP, SBP, DBP, gestational age at delivery, and neonatal weight were analyzed using unpaired t-test. Gestational age at sample, Apgar (1 minute), and Apgar (5 minute) were analyzed with Mann-Whitney *U* test. Values presented as mean with minimum and maximum unless otherwise noted. BMI, body mass index at first obstetric visit; MAP, mean arterial blood pressure at time of blood sample; SBP, systolic blood pressure at time of blood sample; DBP, diastolic blood pressure at time of blood sample; NSVD, normal spontaneous vaginal delivery; VAVD, vacuum-assisted vaginal delivery.

**Table S2**. **Primer, probe, and synthetic standard nucleotide sequences for absolute qPCR of mitochondrial DNA**

| | |
|---|---|
| mtDNA (*MT-ND5*) F: | 5’- GGC ATC AAC CAA CCA CAC CTA -3’ |
| mtDNA (*MT-ND5*) R: | 5’- ATT GTT AAG GTT GTG GAT GAT GGA -3’ |
| TaqMan probe: | 5’- **6FAM** CAT TCC TGC ACA TCT G **MGBNFQ** -3’ |
| SS (gBlock) F: | 5’- TG TTC TGT TCA TTG TTA AGG TTG TGG ATG ATG GAC CCG GAG CAC ATA AAT AGT CGT TAT TTG AAG AAG GCG TGG GTA CAG ATG TGC AGG AAT GCT AGG TGT GGT TGG TTG ATG CCG ATT GGA TTG -3’ |
| SS (gBlock) R: | 5’- CAA TCC AAT CGG CAT CAA CCA ACC ACA CCT AGC ATT CCT GCA CAT CTG TAC CCA CGC CTT CTT CAA ATA ACG ACT ATT TAT GTG CTC CGG GTC CAT CAT CCA CAA CCT TAA CAA TGA ACA GAA CA -3’ |

F, forward. R, reverse. *MT-ND5*, mitochondrial NADH:ubiquinone oxidoreductase core subunit 5. 6FAM, 6-Carboxyfluorescein. MGBNFQ, minor groove binder non-fluorescent quencher. SS, synthetic standard

**Table S3. Variable groupings used for dataset generation.**

| | | |
|---|---|---|
| **Included in all datasets** | **Group 2: Neonatal characteristics** | **Group 4: Extracellular mtDNA signaling** |
| nDNA ng/ml plasma | Neonatal birth length (cm) | mtDNA pg/ml plasma (membrane-bound) |
| DNase1 ng/ml plasma | Neonatal head circumference (cm) | mtDNA pg/ml plasma (membrane-unbound) |
| Mode of delivery | Gestational age at delivery (days) | |
| Maternal age at delivery | Neonatal birth weight (g) | |
| Neonatal sex | Apgar1 score | |
| BMI at NOB | Apgar5 score | |
| **Group 1: Maternal blood pressure** | **Group 3: Maternal reproductive history** | |
| Systolic blood pressure (SBP) | History preeclampsia | |
| Diastolic blood pressure (DBP) | Maternal gravidity | |
| Mean arterial pressure (MAP) | Maternal parity | |
| Chronic hypertension | Number spontaneous abortions | |

Datasets were comprised of select variables included in all datasets and one variable randomly chosen from Groups 1-4. Groupings of related variables determined *a priori*; variables included in all datasets were those that were independent from all other variables in the original dataset. BMI, body mass index; NOB, time of first obstetric appointment.

**Table S4. Coefficients and odds ratios for patient characteristics (elastic net)**

| | Coeff | Coeff SE | OR | OR SE | OR CI (95%) |
|---|---|---|---|---|---|
| Intercept | -2.5034 | 0.1410 | 0.0818 | 0.0184 | (0.0457, 0.1179) |
| nDNA, ng/ml plasma | 0.5032 | 0.0822 | 1.6540 | 0.1110 | (1.4365, 1.8716) |
| mtDNA, pg/ml plasma (membrane-unbound) | -0.0111 | 0.0013 | 0.9890 | 0.00131 | (0.9864, 0.9915) |
| DNase I, ng/ml plasma | 0.1617 | 0.0199 | 1.1755 | 0.0221 | (1.1322, 1.2188) |
| BMI | 0.0208 | 0.0015 | 1.0210 | 0.0015 | (1.018, 1.024) |
| History preeclampsia (Yes) | 0.3710 | 0.0206 | 1.4492 | 0.0281 | (1.3941, 1.5043) |
| Neonatal sex (Male) | 0 | 0.0020 | 1 | 0.0020 | (0.9961, 1.0039) |
| Mode delivery (Vaginal) | -0.0584 | 0.0182 | 0.9433 | 0.0174 | (0.9092, 0.9774) |
| Maternal age | 0.0132 | 0.0025 | 1.0133 | 0.0026 | (1.0083, 1.0183) |
| Birth weight | -0.0001 | 7.55e-06 | 0.9999 | 7.55e-06 | (0.9999, 0.9999) |
| MAP | 0.0222 | 0.0006 | 1.0225 | 0.0006 | (1.0214, 1.0236) |

Values in parentheses indicate reference state for coefficients and odds ratios. Values are the result of 500 bootstraps. BMI, body mass index; CI, confidence interval (95%); Coeff, coefficient; MAP, mean arterial pressure; mtDNA, mitochondrial DNA; nDNA, nuclear DNA; OR, odds ratio; SE, standard error

**Table S5. Model comparison**

| | RMSE | SE | CI (95%) |
|---|---|---|---|
| Ridge | 0.2627 | 0.0179 | (0.2276, 0.2978) |
| LASSO | 0.3200 | 0.0193 | (0.2822, 0.3578) |
| Elastic Net | 0.2438 | 0.0196 | (0.2054, 0.2823) |

RMSE: median root mean square error; SE: standard error; CI: confidence interval (95%)

**Table S6. Model accuracy and receiver operating characteristic**

| | Boot Stat | SE | CI (95%) |
|---|---|---|---|
| Model Accuracy | 1 | 0.0012 | (0.9977, 1.0023) |
| AUC ROC | 1 | 0.0009 | (0.9983, 1.0017) |

Boot stat: bootstrap summary statistic; SE: standard error; CI: confidence interval (95%); AUC ROC: area under curve of the receiver operating characteristic

### Supplementary Figures

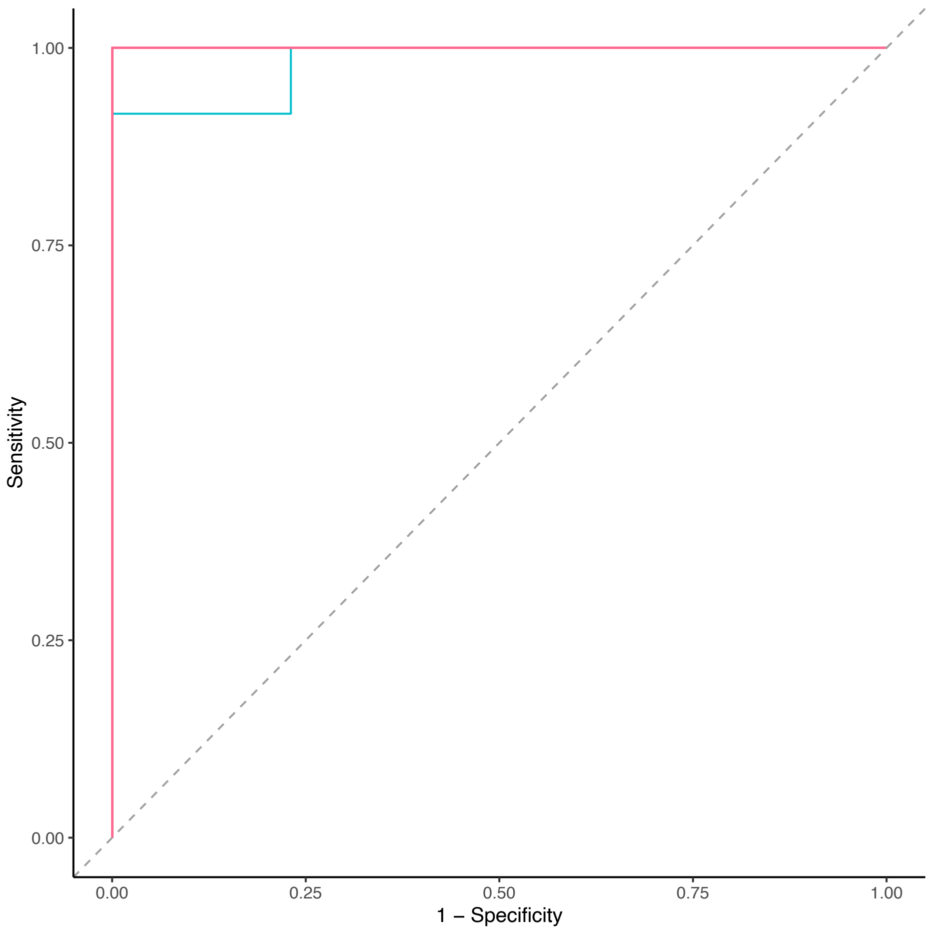


**Figure S1. Accuracy of elastic net penalized regression in the current study.** AUC ROC plot R = 500 simulations (supplementary)
